# Predicting past and future SARS-CoV-2-related sick leave using discrete time Markov modelling

**DOI:** 10.1101/2022.04.22.22274163

**Authors:** Jiayao Lei, Mark Clements, Miriam Elfström, Kalle Conneryd Lundgren, Joakim Dillner

**Affiliations:** Karolinska University Laboratory, Karolinska University Hospital; Department of Medical Epidemiology and Biostatistics, Karolinska Institutet; Karolinska University Hospital

**Keywords:** SARS-CoV-2, COVID-19, Sick leave, Discrete time Markov model

## Abstract

**Background:** Prediction of SARS-CoV-2-induced sick leave among healthcare workers (HCWs) is essential for being able to plan the healthcare response to the epidemic.

**Methods:** During first wave of the SARS-Cov-2 epidemic (April 23^rd^ to June 24^th^, 2020), the HCWs in the greater Stockholm region in Sweden were invited to a study of past or present SARS-CoV-2 infection. We develop a discrete time Markov model using a cohort of 9449 healthcare workers (HCWs) who had complete data on SARS-CoV-2 RNA and antibodies as well as sick leave data for the calendar year 2020. The one-week and standardized longer term transition probabilities of sick leave and the ratios of the standardized probabilities for the baseline covariate distribution were compared with the referent period (an independent period when there were no SARS-CoV-2 infections) in relation to PCR results, serology results and gender.

**Results:** The one-week probabilities of transitioning from healthy to partial sick leave or full sick leave during the outbreak as compared to after the outbreak were highest for healthy HCWs testing positive for large amounts of virus (3.69, (95% confidence interval, CI: 2.44-5.59) and 6.67 (95% CI: 1.58-28.13), respectively). The proportion of all sick leaves attributed to COVID-19 during outbreak was at most 55% (95% CI: 50%-59%).

**Conclusions:** A robust Markov model enabled use of simple SARS-CoV-2 testing data for quantifying past and future COVID-related sick leave among HCWs, which can serve as a basis for planning of healthcare during outbreaks.

## Introduction

Following the emergence of severe acute respiratory syndrome coronavirus 2 (SARS-CoV-2) that caused the coronavirus diseases 2019 (COVID-19), the pandemic resulted in high burden of incidence and mortality worldwide. As of 22 February, 2022, there have been more than 424 million cases and nearly 5.9 million deaths reported (1).

During the pandemic, healthcare workers (HCWs) were highly exposed to SARS-CoV-2 infections and the infections tend to spread rapidly (2). Asymptomatic and pre-symptomatic transmission of SARS-CoV-2 was a major contributor to the COVID-19 pandemic (3, 4). Therefore, detecting and managing healthcare-related SARS-CoV-2 infections timely and effectively are of great significance in preventing onwards transmission, reducing the risk of nosocomial transmission and managing the clusters of COVID-19 (5). Furthermore, prediction of SARS-CoV-2-related sick leave among HCWs is essential for planning of the HCW resources required.

Many countries have routinely tested the HCWs during the pandemic (6–8). At the first wave of the pandemic of COVID-19, the healthcare workers in the Stockholm region, Sweden were offered polymerase chain reaction (PCR) and serological testing to identify the potentially infectious asymptomatic individuals to enable mitigation of the transmission of SARS-CoV-2. PCR testing together with the cycle threshold (Ct) value (the number of cycles required for the fluorescent signal to cross the detectability threshold – a measure of the amount of virus present), provide effective measures to identify infected subjects, also when pre-symptomatic (9). In this study, we aimed to understand the disease burden induced by the infection through modelling the probabilities that healthy HCWs would transition to sick leave in relation to SARS-CoV-2 testing results using a discrete-time Markov model applied to a large-scale HCW cohort in Sweden.

## Material and methods

### Study population

There were approximately 15,300 employees at the Karolinska University Hospital in 2020. During the first wave of the COVID-19 pandemic (between April 23rd and June 24th, 2020), we invited all employees to participate in our SARS-CoV-2 screening study to evaluate the probability of sick leave in relation to presence of viral nucleic acids in throat swabs and presence of antibodies to the virus in serum. All enrolled individuals provided a written informed consent that gave permission to link to the hospital administrative databases to obtain information from sick leave records. We excluded individuals that had missing or invalid testing results, had incomplete consent forms or were not formally employed at the hospital. In the final analysis, we only included individuals that had complete sick leave, PCR testing results and serology results. Among the approximately 15,300 employees at Karolinska University Hospital, a total of 14,201 were enrolled in this study. The cohort that met all inclusion criteria consisted of 9449 individuals. A majority (79.2%) of the individuals were female, PCR negative (97.5%), and serology negative (89.5%).

The study was approved by the National Ethical Review Agency of Sweden (Dnr: 2020-01620). Trial registration number: ClinicalTrials.gov NCT04411576.

### Laboratory test

The viral nucleic acid detection was based on throat swab samples and all the diagnostic procedures followed standard operating protocol validated for reproducibility, sensitivity and specificity, including lack of cross-reactivity with other Coronavirus strains. The SARS-CoV-2 antibody test used an assay with a sensitivity of 99.4% and a specificity of 99.1% (10). The testing methods for both viral nucleic acid and serological results for antibody were described in detailed in our previous publication (10).

### Sick leave

We obtained the sick leave information for each participant starting from 14 weeks before the week of sampling to 28 weeks after the week of sampling from the administrative database of the Karolinska University Hospital. According to the regulations of the Hospital, all employees with symptoms should not have been at work. As HCWs were enrolled at work, they were assumed to be healthy at day 0 at the week of sampling. Employees working from home that developed COVID-19 symptoms were required to report this as sick leave according to the hospital’s rules, which substantially improved the accuracy of the sick leave data. In this study, only sick leave data between calendar week 17 to week 27 were used because the screening tests were performed from week 17 onward, the first wave of excess sick leave for all employees at the hospital diminished by around week 27 comparing the sick leave data in year 2020 to year 2019 (Supplementary figure S1) and an effective lead time between testing results and its impact on sick leave.

### Statistical analyses

PCR results was defined as being strongly positive with Ct value below 27.0, weakly positive with Ct value between 27.0 and 35.2, or negative. The serological results were classified as positive or negative. Sick leave was categorized in three states in each calendar week based on the number of days the individual was on sick leave. More specifically, individuals were classified as either healthy (with 0 days of sick leave), on partial sick leave (with 1 to 4 days of sick leave) or on full sick leave (with 5 to 7 days of sick leave).

Further, we computed the frequency of the transitions across the three sick leave states from the current calendar week to the following calendar week. Multinomial logistic regression models were used to examine the association between the factors including serological results, PCR results, gender, and calendar time in relation to the sick leave state given the previous week’s sick leave state (that is, three regression models). All covariates were mutually adjusted in the regression models. We estimated odds ratios and 95% confidence intervals (CIs). We performed the analysis for the time period after the week of sampling, stratifying by the initial state of healthy, partial sick leave and full sick leave, respectively. Calendar time was modelled using natural splines with one degrees of freedom for those healthy, on partial sick leave and those of full sick leave, respectively.

Based on the regression models, we predicted the one-week transition probabilities (defined as the transitions from the current calendar week to the next calendar week) between the sick leave states. Assuming that individuals were healthy at week 17, we also computed the transition probabilities from week 17 to subsequent weeks using the Chapman-Kolmogorov equation (Technical details in supplementary). We further calculated point estimates, variances and CIs for (a) the sum of the standardized transition probabilities for partial and full sick leave and (b) the ratio of the standardized transition probabilities for the baseline covariate distribution compared with the referent period (an independent period from week 28 to week 34, assuming low circulation of SARS-CoV-2), together with sick leave that attributable to COVID-19. These predictions for transition probabilities were varied by PCR, serology and gender. Variance estimates for these transition probabilities were calculated using the Delta method, where the partial derivatives of the transition probabilities with respect to the regression parameters were calculated using finite differences, and where the full covariance matrix from the three regression models was assumed to be block-diagonal. The 95% CIs for the transition probabilities were computed using the Delta method on a logit scale. We used R software, version 4.0.5 (R Foundation for Statistical Computing) for data management and statistical analyses.

## Sensitivity analyses

As a sensitivity analysis, we varied the multinomial logistic regression models, with additional adjustments for age groups and contact with patients to ensure that the adjustments in our main models were robust. Moreover, we used an extended period of the sick leave data from week 17 to week 44 in order to examine the internal validity of our results.

## Results

### SARS-CoV-2 tests and risk of sick leave in the next week

Among individuals with a healthy state (being at work and not on sick leave) in the preceding week (Table 1a), having a positive serology was associated with a significantly decreased risk of being on partial sick leave (sick leave 1-4 days in the next week) compared to having negative serology (odds ratio (OR) 0.74, 95% CI: 0.63-0.87). Individuals with strongly positive PCR results were associated with a significantly increased risk of being in partial sick leave with an OR of 1.75, (95% CI: 1.12-2.74) compared to PCR negative individuals, while individuals with weakly positive PCR results did not have a statistically significant increased risk for partial sick leave in the next week (Table 1a). For individuals on partial sick leave in the preceding week (Table 1b), those having a strongly positive PCR were associated with significantly increased risk of being on full sick leave for the entire next week (2.48, (95% CI: 1.41-4.34)). In this context, it should be noted that the entire study was performed at a time when there was no ongoing testing and the testing performed by the research group took two weeks, thus the testing result itself did not influence the probability for sick leave in the next week.

**Table 1.** Odds ratios of sick leave state after the week of sampling up to calendar week 27.

**a) Initial state: healthy**
|  | **To sick leave** |  |
| --- | --- | --- |
|  | **Partial sick leave** | **Full sick leave** |
| **Serological results** |  |  |
| Positive vs. Negative | 0.74 (0.63-0.87) | 0.81 (0.42-1.53) |
| **PCR results** |  |  |
| Strongly positive vs. Negative | 1.75 (1.12-2.74) | 3.19 (0.78-13.00) |
| Weakly positive vs. Negative | 0.80 (0.54-1.17) | 1.57 (0.48-5.14) |
| **Gender** |  |  |
| Male vs Female | 0.50 (0.44-0.57) | 0.65 (0.40-1.07) |

**b) Initial state: partial sick leave**
|  | **To sick leave** |  |
| --- | --- | --- |
|  | **Healthy** | **Full sick leave** |
| **Serological results** |  |  |
| Positive vs. Negative | 1.13 (0.88-1.46) | 0.96 (0.62-1.47) |
| **PCR results** |  |  |
| Strongly positive vs. Negative | 0.69 (0.42-1.12) | 2.48 (1.41-4.34) |
| Weakly positive vs. Negative | 1.01 (0.62-1.63) | 0.87 (0.37-2.04) |
| **Gender** |  |  |
| Male vs Female | 1.29 (1.04-1.59) | 0.95 (0.66-1.37) |

**c) Initial state: full sick leave**
|  | **To sick leave** |  |
| --- | --- | --- |
|  | **Healthy** | **Partial sick leave** |
| **Serological results** |  |  |
| Positive vs. Negative | 1.69 (0.65-4.40) | 0.99 (0.45-2.17) |
| **PCR results** |  |  |
| Strongly positive vs. Negative | 0.24 (0.05-1.10) | 0.66 (0.31-1.41) |
| Weakly positive vs. Negative | 1.06 (0.19-5.91) | 0.90 (0.22-3.65) |
| **Gender** |  |  |
| Male vs Female | 1.06 (0.46-2.45) | 0.91 (0.49-1.69) |

- Adjust for PCR, serology results, gender and calendar period with one degree of freedom.

### SARS-CoV-2 testing and transition probability of sick leave in relation to calendar time

During the calendar week 17 to week 27, the one-week transition probabilities from healthy to partial sick leave or full sick leave showed a decreasing trend over time (Figure 1), and were higher among individuals who had a strongly positive PCR, had negative serology, and were females. No major differences over time were found for the one-week transition probabilities for individuals with either partial sick leave or full sick leave in the preceding week (Supplementary figure S2).

**Figure 1.**
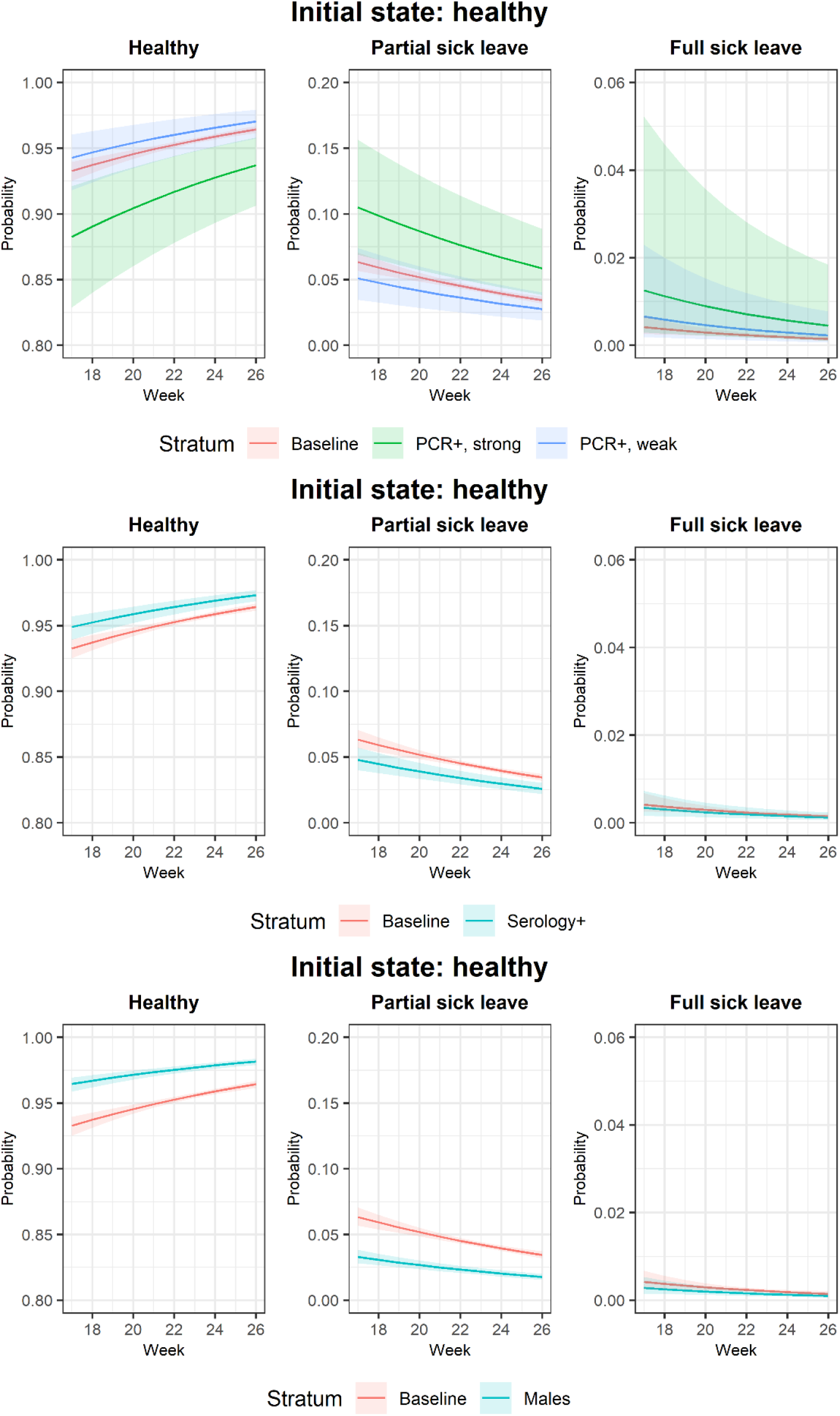
Predicted one-week transition probabilities from healthy state by calendar week.

The sum of the standardized transition probabilities for sick leave in the upcoming week for those in the healthy state at week 17 with a strongly positive PCR results decreased from 100% to slightly above 80% (Figure 2). These probabilities stayed above 90% over the period for the rest of the group. The standardized transition probabilities of being on partial sick leave increased to almost 15% for individuals with strongly positive PCR, while for the individuals with positive serology, these probabilities only increased to around 5%. As for full sick leave, we observed very low probabilities for all groups except for individuals with strongly positive PCR: around 5% of individuals were on full sick leave also from week 20.

**Figure 2.**
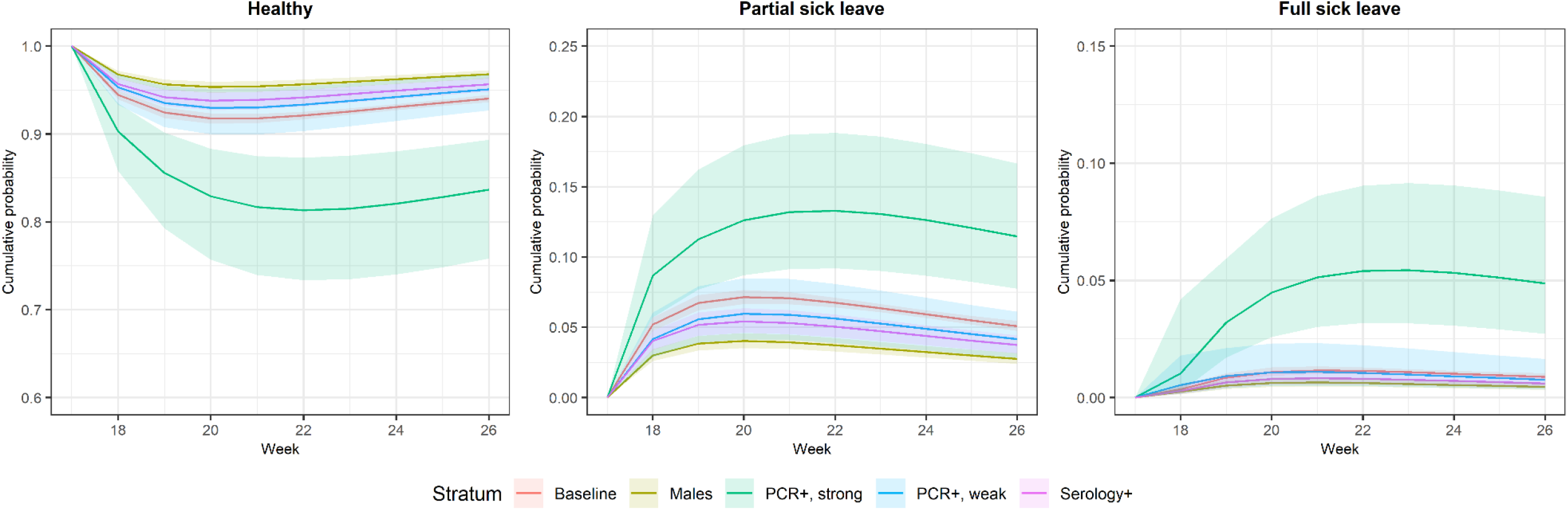
Standardized transition probabilities by calendar week.

### Attributable fraction of sick leave due to SARS-CoV-2

Compared to the reference period with no or low circulation of SARS-CoV-2, the ratios of transition probabilities were highest for individuals with strongly positive PCR for either partial sick leave or full sick leave (Figure 3). The ratios peaked at week 18 for transitioning to partial sick leave 3.69 (95% CI: 2.44-5.59) and to full sick leave 6.67 (95% CI: 1.58-28.13) compared to the reference period. During the study period, the proportion of all sick leave that could be attributed to SARS-CoV-2 was estimated to be 55%, (95% CI: 50%-59%) at week 18, and decreased to 16% (95% CI: 7%-23%) by week 26 (Figure 4 and Supplementary figure S3).

**Figure 3.**
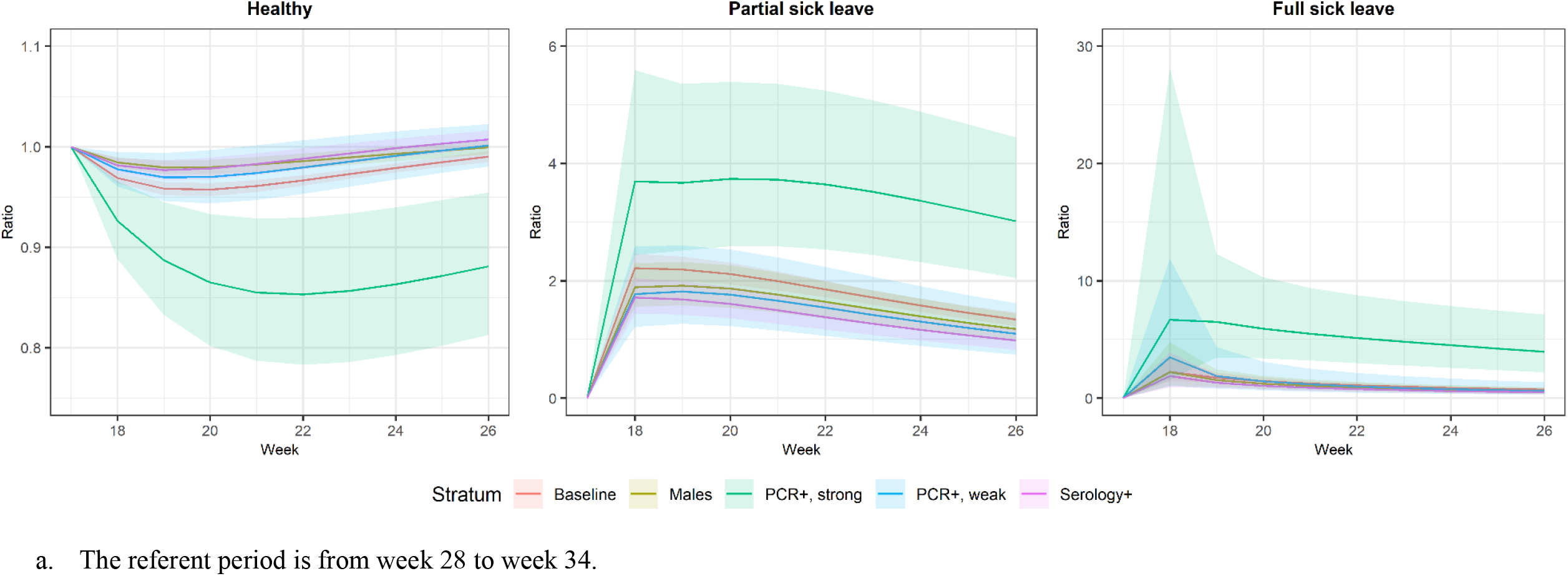
Ratios of longer-term standardized transition probabilities comparing to referent period by calendar week.

**Figure 4.**
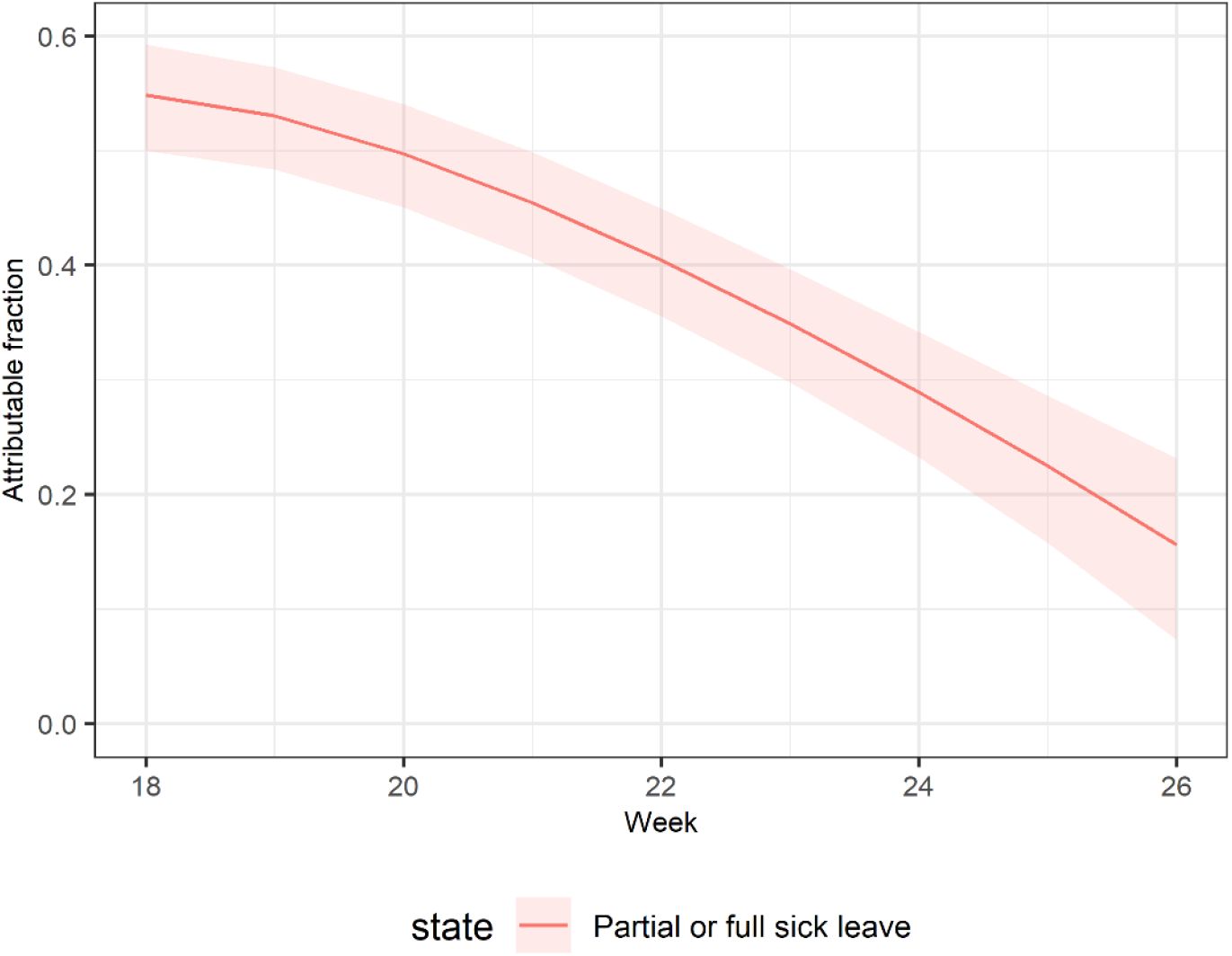
Attributable fraction of sick leave due to COVID-19 by calendar week.

### Sensitivity analyses

By including additional covariates of age group and contact with patients, the results from the multinomial logistic regression were comparable with the estimates from the main analysis (Supplementary table S1-S2). Using all available sick leave data from the week of sampling gave similar results when estimating the ORs for sick leave (Table S3-S4). In addition, the predicted one-week transition probabilities from calendar week 17 to week 44 and standardized cumulative transition probabilities showed consistent tends in comparison to the period from week 17 to 27. We observed a second peak of the transition probabilities around week 38 consistent with the second wave of SARS-CoV-2 infections in Sweden (Supplementary Figure S4 – S5).

## Discussion

### Interpretation of results

In this large-scale cohort study, we found that the strongly PCR positive asymptomatic HCWs had an increased risk of and higher transition probabilities to sick leave in the next week. The predicted one-week transition probabilities from healthy to either of the sick leave states declined as the outbreak subsided. The proportion of all sick leaves that could be attributed to SARS-CoV-2 declined from 55% to 16% during the study period. As individuals with symptoms should not be at work according to the hospital regulations, the PCR positive individuals in our study will generally represent pre-symptomatic, asymptomatic, or post-symptomatic infection. We have previously described that low amount of virus is indicative of post-symptomatic infections (previously symptomatic who have returned to work after becoming healthy) and high amounts of virus indicates pre-symptomatic infection (10). There was also a large proportion of individuals at work who with strongly positive PCR who remained in the healthy state (10). In this study, we have performed a formalized quantitation of the future sick leave risk in relation to simple SARS-CoV-2 testing results, which may be useful for understanding the disease-inducing propensity of the infection and for planning of the supply of HCW personnel needed in the upcoming weeks.

In contrast, being serology positive showed a very low predicted longer-term transition probabilities of being in either partial sick leave or full sick leave state (much less than serology negatives), which indicates immunity. While there was not a major difference in terms of gender or serology results for the predicted longer-term transitioning probabilities, being PCR positive, especially being strongly PCR positive, suggested a pronounced probability of being on future sick leave. A major proportion of transitioning from healthy to sick leave occurred within the first week, in line with the reported serial interval of SARS-CoV-2 of around 6 days (4).

The changes of both next week and longer-term transition probabilities predicted from our model together with sick leave that attributed to COVID-19 reflect the circulation of infections of SARS-CoV-2 among the population during the study period: the outbreak peaked at the first quarter of 2020 and starting to decline afterwards until mid-2020. This is well in line with the reported burden of COVID-19 during this time period in Sweden (11). It should be noted that there is a strong seasonality of the likelihood for sick leave, presumably mostly caused by seasonal circulation of other infections.

The risk for HCWs to acquire infections with SARS-CoV-2 and to become sick does not imply that the infections necessarily occurred in the healthcare setting, as there was ongoing transmission in the community (7,12). The fact that the SARS-CoV-2 test results during the first outbreak in the spring of 2020 did predict sick leave also in the second outbreak around week 38 should be interpreted with caution as knowledge of the test results could have influenced behaviour.

### Comparison with other studies

The proportion of PCR positivity detected in asymptomatic individuals was similar to a HCW study based in London during a similar period (8). A HCWs study conducted in Spain also suggested that COVID-19 caused long periods of sick leave (7). We also confirmed the previous findings that the seropositive individuals have a decreased risk for future sick leave (13). Many studies have suggested that pre-symptomatic or asymptomatic infections might drive the transmission (5,14), which is in line with our results on standardized longer-term transition probabilities for the individuals with strongly positive PCR results. We have not been able to locate other cohort studies linking SARS-CoV-2 testing results to sick leave data.

### Strength and limitations

We used routinely collected sick leave data over a long period based on the administrative information from a large and systematically enrolled hospital HCW cohort, minimizing recall bias and ensuring high data quality. Using information from the administrative database in the hospital allowed us to have virtually complete information on sick leave and therefore increased the validity of our findings. The serology platform used in our study contained several SARS-CoV-2 proteins and was validated for high sensitivity and specificity. We have also been able to show the one-week and longer-term transition probabilities from one state to another state over the study period as well as attributable fraction of sick leave due to COVID-19, which adds the understanding on impact of infections of SARS-CoV-2 on sick leave beyond the previous evidence. Moreover, we have provided a novel example of using discrete-time Markov models for analyzing such data that is not commonly used in the current literature (15).

There were some limitations in our study. First, we have only been able to measure the infections of SARS-CoV-2 through PCR and serology at the baseline instead of repeated measurement to identify the diseases states. However, we have followed up sick leave data which was a less biased measures for future infections and occurrence of diseases. Second, we were not able to test the HCWs that were not at work which might include a proportion of individuals that were infected by SARS-CoV-2. Third, even though the comparison with empirical sick leave suggested that the level of sick leave during the reference period was comparable to the same period in 2019, a small magnitude of sick leave recorded in this period could still be COVID-19 related. If so, we may have underestimated the attributable fraction to some extent.

In conclusion, the probability for asymptomatic individuals at work to transition to sick leave could be robustly estimated with a Markov model based on ordinary SARS-CoV-2 testing data. A high and quantifiable proportion of all sick leave during the pandemic could be attributed to infection with SARS-CoV-2. The factor that was most important for sick leave prediction was presence of high amounts of virus, implying that the Ct values of the PCR testing could be important for the planning of required healthcare personnel resources in response to the SARS-CoV-2 epidemic.

## Data Availability

The data constitutes sensitive data about health of human research subjects and thus cannot be directly deposited openly. However, pseudonymised, individual-level data that allow full replication of the results in this article are available from the corresponding author on reasonable request.

## Declarations

### Ethics approval and consent to participate

The study was approved by the National Ethical Review Agency of Sweden (Dnr: 2020-01620).

### Authors’ contributions

JD and KCL conceived the research questions and hypotheses; MC developed the model; ME conducted the data management for the initial cohort; JL and MC performed the analysis; JL and MC wrote the original draft of this manuscript; All authors interpreted the data, involved in the critical revision of article and have read and approved submission of the manuscript.

## Acknowledgements

We would like to thank Suyesh Amatya, Helena Andersson, Shaghayegh Bayati, Emine Eken, Pedram Farsi, Yasmin Hussein, Roxana Merino Martinez, Sara Mravinacova, Björn Pfeifer, Ulla Rudsander, Sadaf Sakina Hassan, Ronald Sjöberg, Balazs Szakos, Hanna Tegel, and Emel Yilmaz for excellent technical assistance.

